# Significant Structural Breaks in the International Incidence of Hepatitis Delta Virus

**DOI:** 10.1101/2021.07.12.21260388

**Authors:** Braden S. Fallon, Elaine M. Cooke, Matthew C. Hesterman, Jared S. Norseth, Sherzod B. Akhundjanov, Melodie L. Weller

## Abstract

**Background & Aims:** The international incidence of Hepatitis Delta Virus (HDV) is challenging to accurately estimate due to limited testing and lack of active surveillance for this rare infectious disease. These limitations prevent the detection of low-level and/or geographically dispersed changes in the incidence of HDV diagnoses. A study was designed to enable international active tracking and analyses of HDV epidemiology by aggregating international HDV and Hepatitis B Virus (HBV) diagnoses datasets.

**Methods:** Publicly accessible datasets containing yearly incidence for HDV and HBV diagnoses were mined from government publications for Argentina, Australia, Austria, Brazil, Bulgaria, Canada, Finland, Germany, Macao, Netherlands, New Zealand, Norway, Sweden, Taiwan, Thailand, United Kingdom, and United States. The Bayesian Information Criterion (BIC) was used to determine the best-fitting breakpoint model for the number of breakpoints and the break dates identified using the determined model.

**Results:** Aggregated analysis of these HDV and HBV datasets spanning 1999-2020 identified structural breaks in the timeline of HDV incidence in 2002, 2012, and 2017. A significant increase in the international HDV incidence, relative to reported HBV diagnoses, occurred in 2013-2017. Secondary analysis identified four distinct temporal clusters of HDV incidence, including Cluster I (Macao, Taiwan), Cluster II (Argentina, Brazil, Germany, Thailand), Cluster III (Bulgaria, Netherlands, New Zealand, United Kingdom, United States), and Cluster IV (Australia, Austria, Canada, Finland, Norway, Sweden).

**Conclusion:** Re-evaluation of the testing paradigm for HDV in HBV-positive patients and an active surveillance status of HDV are warranted to define the etiology of the structural breaks in HDV incidence timelines.

## INTRODUCTION

Worldwide, it is estimated that 12-72 million individuals are infected with Hepatitis Delta Virus (HDV)^1–4^. Regionally, the prevalence of HDV in Hepatitis B Virus (HBV) positive populations can range from less than 1% in North America and Europe to greater than 20% in parts of South America, Asia, and Africa. The accuracy of these prior estimates is heavily influenced by the lack of active surveillance of international HDV diagnoses and limited testing in at-risk populations^5–7^. Recent reports have alluded to a provocative shift in HDV epidemiology, including the identification of new at-risk populations and HDV-like sequences in diverse animal species^8–15^. Improved infectious disease reporting networks and education are essential to track critical changes in HDV epidemiology, including the emergence of new HDV variants, alternative reservoirs, and novel transmission patterns of this rare infectious disease.

Since the discovery of HDV in the late 1970s, the molecular characteristics of this unique pathogen have been extensively studied^16^. HDV is a satellite RNA that requires a helper virus, classically defined as HBV, for packaging and transmission^17^. As one of the smallest viruses known to infect humans, HDV contains a ∼1700nt single-stranded, circular RNA genome that supports expression of two antigens from a single open reading frame. The HDV genome and two antigens comprise the ribonucleoprotein complex that is packaged into the HBV membrane. The tropism of HDV is dictated by the tropism of the helper virus. Once intracellular, HDV utilizes the host cellular machinery for replication and can persist chronically in the absence of an active helper virus co-infection^17–19^.

Our understanding of HDV is still rapidly evolving. Hepatitis Delta Virus is a member of the new Kolmioviridae family^20^ that includes 8 new genus and HDV-like sequences isolated from birds, bats, deer, rodents, snakes, newts, toads, and termites^9–14^. A majority of these new HDV-like variants were detected in the absence of a Hepadnaviridae coinfection. Additionally, HDV has been detected in non-hepatic tissue, including the salivary glands of patients with primary Sjogren’s syndrome (pSS), and in the absence of a detectible past or current HBV co-infection^8^.

The absence of active surveillance and limited testing for HDV impedes the ability to accurately detect changes in the international HDV epidemiology. To address this, a study was designed to identify publicly accessible infectious disease datasets from around the world to enable collective tracking and analyses of international HDV incidence. To date, 17 international infectious disease datasets containing yearly incidence of HDV and HBV that span five continents have been identified. Analyses of these datasets have identified significant structural breaks in the HDV diagnoses timelines between 1999-2020, including four distinct temporal clusters that are suggestive of significant changes in the HDV incidence profiles.

## METHODS

### International HDV Public Datasets and Analyses

International infectious disease datasets containing yearly diagnosis information for both HDV and HBV were obtained through data mining techniques. These data mining techniques included searching for governmental infectious disease datasets and publications through localized, language-specific methods, and geographically distinct search engines. All infectious disease data sources identified are noted in Supplemental Table 1. Publicly accessible infectious disease datasets for 16 countries and one administrative region containing both yearly incidence of HDV and HBV include Argentina, Australia, Austria, Brazil, Bulgaria, Canada (BC), Finland, Germany, Netherlands, New Zealand, Norway, Sweden, Taiwan, Thailand, United Kingdom, United States, and Macao. Datasets contain the number of HDV and HBV diagnoses that occur per year. Limited information is available on the number of tests performed, type of HDV and HBV testing used, patient-matched HBV and HDV testing results or other regulations impacting the reporting of HDV and HBV diagnoses. Details of the HBV and HDV testing paradigm for the United States’ Center for Disease Control (CDC) National Health and Nutritional Examination Survey (NHANES) are detailed in the supplemental methods and Supplemental Figure 1. Current HDV diagnostic paradigms restrict testing for HDV to patients with documented acute or chronic HBV infections. Prevalence of HDV was calculated as the percent of total HDV diagnoses per total HBV diagnoses or total HDV diagnoses per 100,000 HBV diagnoses. Yearly and cluster HDV incidences were calculated as the ratio of yearly HDV diagnoses to the total yearly HBV diagnoses identified from each dataset and reported as the average of yearly HDV diagnoses/100,000 HBV diagnoses (HDV/HBV_100,000_) with 95% confidence interval (CI). Pairwise correlations of yearly HDV/HBV incidences for each country relative to yearly incidence of all other countries were calculated using an unconditional Pearson correlation coefficient. Correlative heatmap with hierarchal clustering and a temporal heatmap of maximal HDV/HBV yearly incidence were produced using Morpheus (Broad Institute, https://software.broadinstitute.org/morpheus/).

### Structural Break Analysis

To identify structural shifts in the HDV/HBV time series, a structural break analysis was performed as detailed by Bai and Perron^21,22^ and as implemented by Zeileis, et al^23^. This methodology has been utilized prior to evaluate datasets for structural breakpoints in infectious disease time series^24–26^. The advantage of the Bai and Perron method is that it allows for endogenous identification of single or multiple breakpoints in the dataset timeline. Breakpoints in the mean of HDV/HBV series, which is of primary interest, were determined by fitting a constant model to the data and analyzing level structural changes^22,23,27^. The null hypothesis model of no structural break was tested against a set of alternative hypothesis models of 1, 2, or more structural breaks. The Bayesian Information Criterion (BIC) was used to determine the best-fitting breakpoint model for the number of breakpoints and the break dates identified using the determined model. Using data from the correlation matrix, four country clusters were identified. Structural break analysis was performed on both the aggregated dataset and in cluster datasets identified through correlative hierarchal clustering as previously detailed.

### Statistical Analysis

Statistical analyses, including standard student t-test, were performed on all available HDV/HBV incidence for the identified structural breaks in the HDV/HBV timelines and subsequent cluster data.

## RESULTS

### International HDV and HBV Prevalence

Seventeen publicly accessible infectious disease datasets containing the yearly incidence of HDV and HBV diagnoses were identified and utilized in this study. The source data for each publicly accessible dataset are contained in Supplemental Table 1. Infectious disease datasets containing the yearly incidence of HDV and HBV diagnosis were located for Argentina, Australia, Austria, Brazil, Bulgaria, Canada, Finland, Germany, Macao, Netherlands, New Zealand, Norway, Sweden, Thailand, Taiwan, United Kingdom, and United States. Analyses were conducted on complete and partial datasets ranging between 1999-2020. Together, these aggregated datasets contain >700,000 HBV and >9,000 HDV diagnoses that spans 5 continents and represents one of the largest active HDV and HBV datasets analyzed.

The prevalence of HDV diagnoses relative to HBV diagnoses was calculated for all 17 datasets for the publicly available time frames identified (Table 1). Prior studies have reported the prevalence of HDV in HBV-positive patient populations to be approximately 5% with areas of lower or higher prevalence^1^. From our analysis, the average prevalence of HDV across the identified infectious disease datasets is 2·56% (95% CI 0·18%-4·94%). Percent prevalence of HDV diagnoses relative to HBV diagnoses ranged from 0·26% in Canada to 20% in the United States (1999-2018, NHANES). Yearly incidence of HDV diagnoses relative to HBV diagnoses were calculated for each of the 17 datasets (Figure 1). A trend of increasing yearly incidence in HDV diagnoses relative to HBV diagnoses was observed in a subset of the datasets analyzed. Further analyses were performed to evaluate the presence of structural breaks in the HDV/HBV incidence time series and international correlations within these HDV/HBV incidence timelines.

**Table 1.**
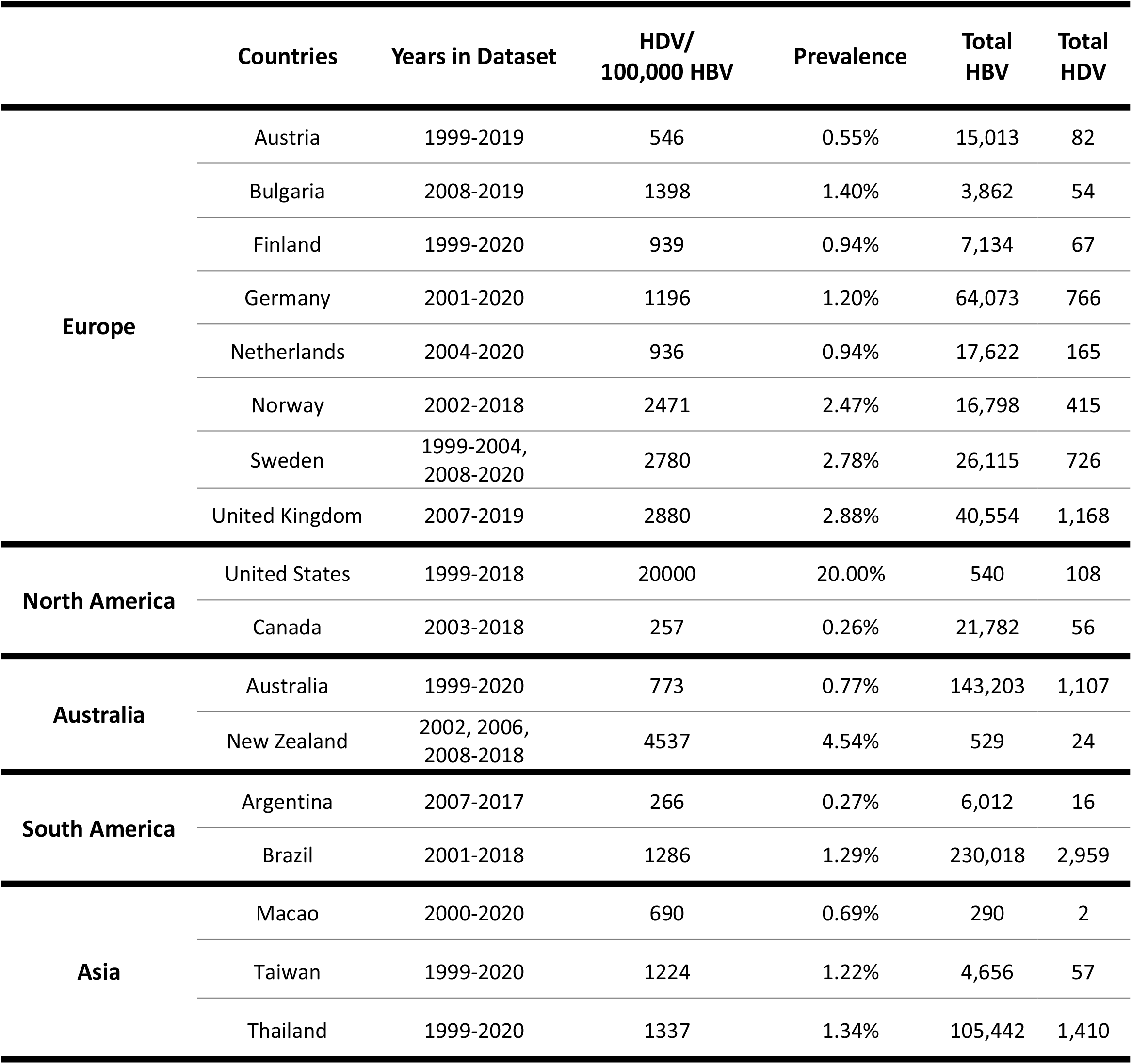
Prevalence of Hepatitis Delta Virus for infectious Disease Datasets Identified. Publicly accessible epidemiological datasets from 17 countries were identified that contained both the yearly incidence of HDV and HBV diagnoses.

**Figure 1.**
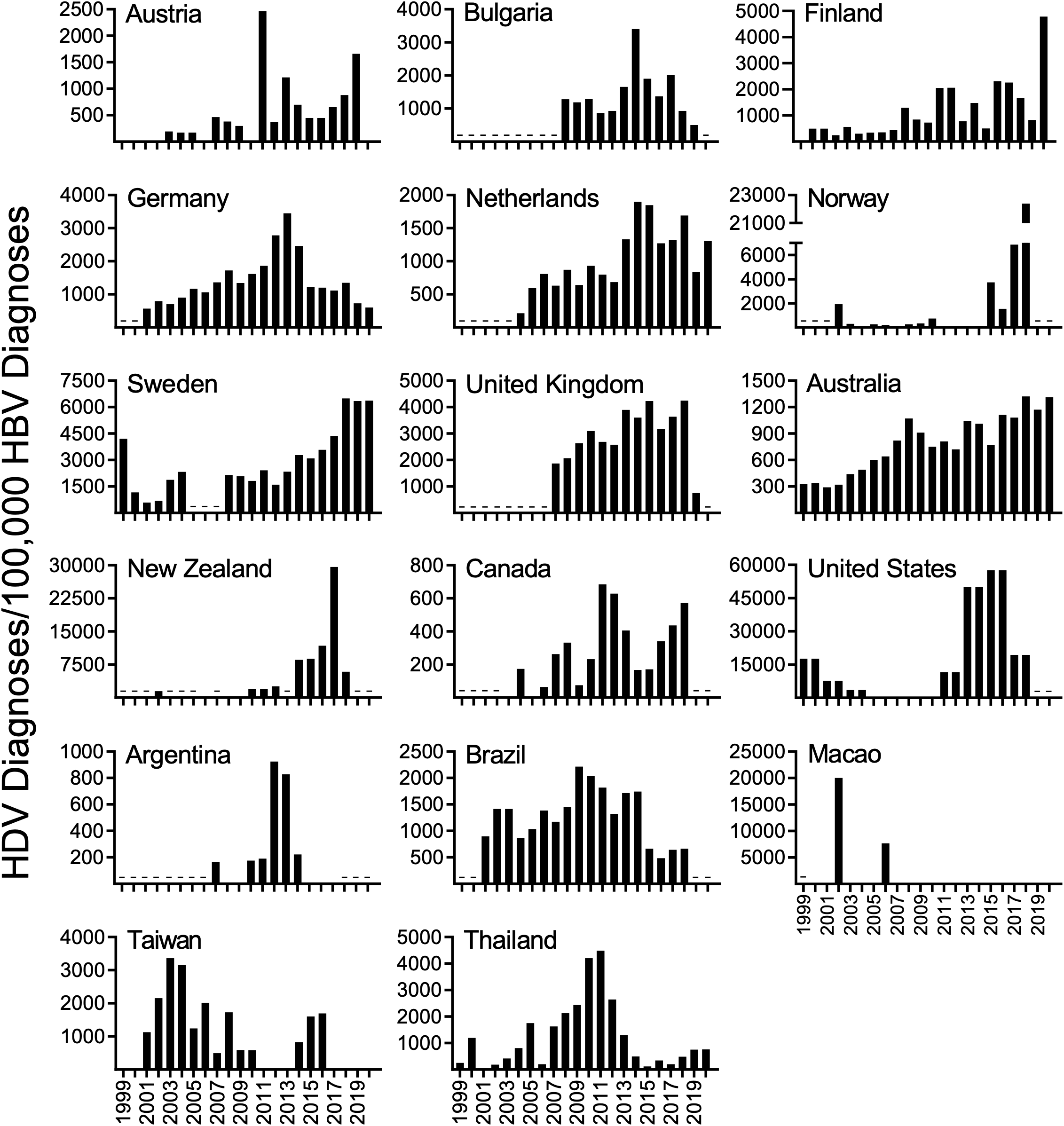
Yearly incidence of HDV per 100,000 HBV diagnoses. Publicly accessible infectious disease datasets containing yearly incidence of HDV and HBV for 17 countries spanning 5 continents were utilized in analyses. Data ranges in full or in part between 1999-2020. Data not accessible (-).

### Structural breaks in aggregated HDV incidence occurred in 2002, 2012, and 2017

Hepatitis Delta Virus (HDV) transmission is predominantly associated with parenteral, sexual, contaminated blood or blood products, and vertical exposure as a co-infection with HBV, or a superinfection in chronic HBV infected individuals^1,28,29^. Therefore, given the known routes of HDV transmission, the shared trend of increasing yearly incidence of HDV spanning multiple continents was not expected and warranted further evaluation. Breakpoint analysis was performed to identify the presence and timeline of structural breaks in the incidence of HDV relative to reported HBV diagnoses. The HDV/HBV time series was fitted using a constant model with no structural break (null hypothesis model) and compared against a model with 1, 2, or more structural breaks (alternative hypothesis model). The Bayesian Information Criterion (BIC) was used to identify the presence and number of potential breaks in the time series. Through this analysis, it was found that a model with 3 breakpoints represented the lowest BIC value and therefore was the best fitting model (Supplemental Figure 2). Using this breakpoint analysis, the break dates in the aggregate HDV incidence data were identified as 2002, 2012, and 2017 with accompanying 95% confidence intervals (Figure 2).

**Figure 2.**
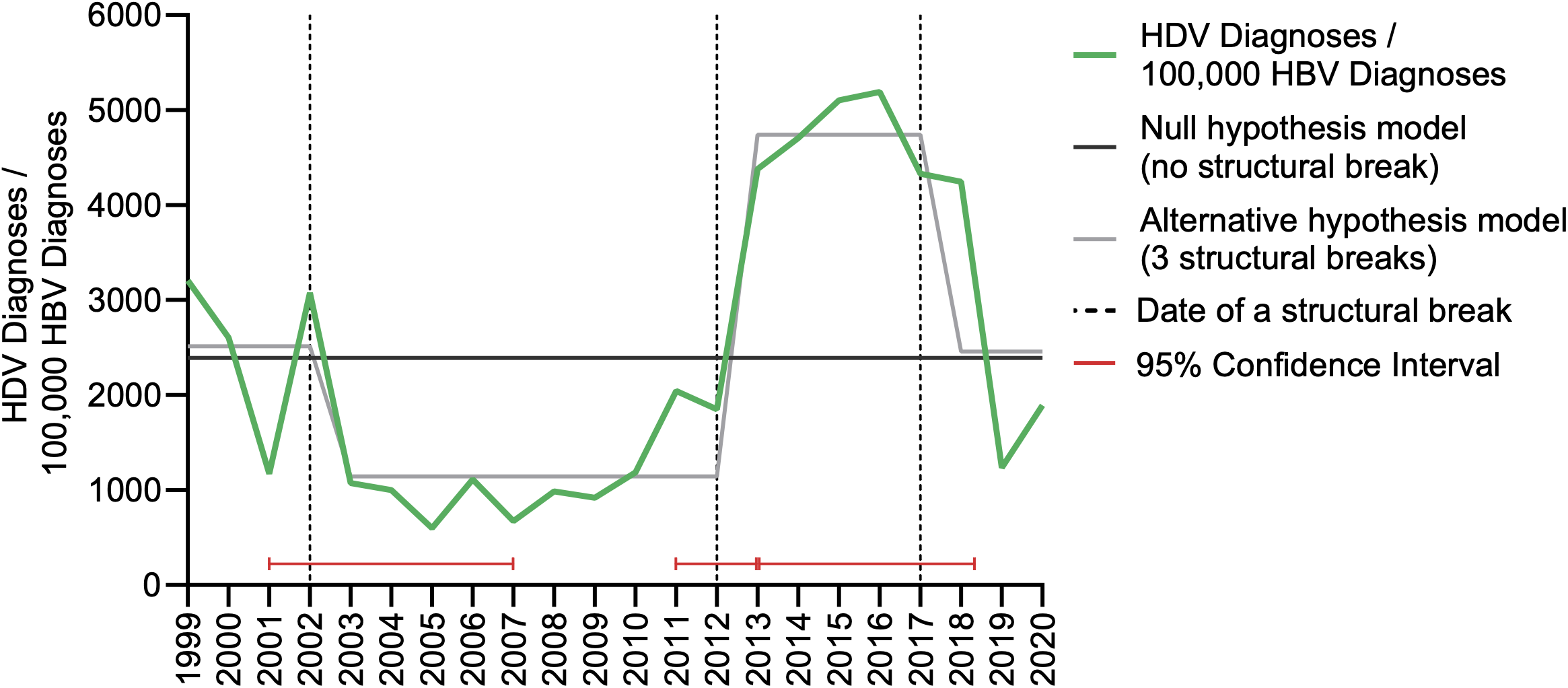
Structural Break Analysis identified three structural breaks at 2002, 2012 and 2017 in the aggregated yearly incidence of HDV diagnoses/100,000 HBV Diagnoses. The presence of three structural breaks in average HDV diagnoses/100,000 HBV diagnoses occur in 2002, 2012 and 2017. Fitting the null hypothesis model (no structural break) and alternative hypothesis model (3 structural breaks) to the data indicate clear structural shifts at those times.

For control measures and robustness, one additional breakpoint analysis was performed. This analysis utilized the aggregate HDV/HBV data for all countries except the United States (exclusion of NHANES dataset). NHANES HBV and HDV datasets are acquired through a general surveillance program where HBV-positive samples undergo automatic reflex testing for HDV, and HBV and HDV testing is initiated regardless of hepatic symptomology of infection. Therefore, additional structural break analyses were performed without NHANES to evaluate for potential bias introduced by the alternative testing protocols. The additional breakpoint analysis identified a single structural break in the HDV/HBV time series after 2013 with accompanying 95% confidence interval (Supplemental Figure 3).

### Significant change in HDV incidence for identified breakpoints

Aggregated incidence of HDV was evaluated based on the three identified structural breakpoints by BIC. Based on these breakpoints, the average yearly incidence of HDV significantly decreased between the periods of 1999-2002 (2,515 HDV/HBV_100,000_) and 2003-2012 (1,144 HDV/HBV_100,000_; Fold Change (FC):-2·2; p=0·03), increased between the periods of 2003-2012 (1,144 HDV/HBV_100,000_) and 2013-2017 (4,741 HDV/HBV_100,000_; FC:4·14; p<0·0001) and decreased again between the periods of 2013-2017 (4,741 HDV/HBV_100,000_) and 2018-2020 (2,458 HDV/HBV_100,000_; FC:-1·93; p=0·003) (Figure 3A).

**Figure 3.**
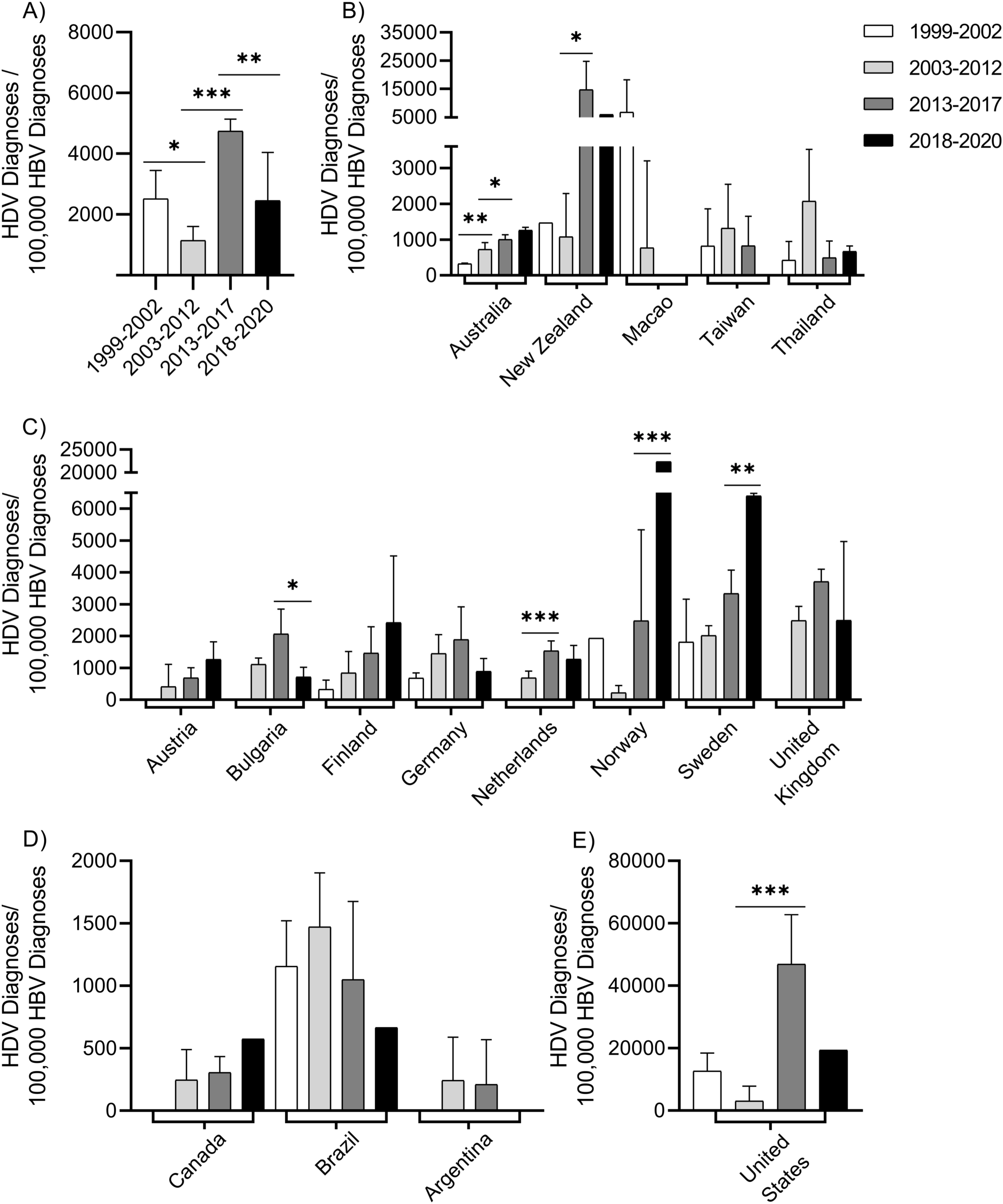
HDV/HBV incidence and country-level HDV diagnoses per 100,000 HBV diagnoses compared for 1999-2002, 2003-2012, 2013-2017 and 2018-2020. A) Comparison of HDV/HBV incidence for identified breakpoints in the aggregated data, Country level analyses for B) Asia and Australia, C) Europe, D) Americas, and E) United States. *p ≤ 0.05, **p ≤ 0.01, ***p ≤ 0.001

Of interest are seven countries that significantly go against the global HDV incidence trends. Six countries have significant increases in HDV incidence at time points where global trends are declining. These countries include Australia, New Zealand, Netherlands, Norway, Sweden, and the United States (Figure 3 B-E). Australia had a significant increase in HDV incidence from the time period 1999-2002 to 2003-2012 (321 vs 726 HDV/HBV_100,000_; p=0·0014). Australia, Netherlands, New Zealand, and the United States all experienced a significant increase in HDV incidence from the time period of 2003-2012 to 2013-2017 (Australia: 726 vs 1003 HDV/HBV_100,000_; p=0·017; Netherlands: 686 vs 1,535 HDV/HBV_100,000_; p=0·0003; New Zealand: 1,081 vs 14,698 HDV/HBV_100,000_; p=0·039; United States: 3,040 vs 46,901 HDV/HBV_100,000_; p<0·0001). Sweden and Norway experienced a significant increase in HDV incidence from the time period 2013-2017 to 2018-2020 (Norway: 2,481 vs 22,398 HDV/HBV_100,000_; p<0·0001; Sweden: 3,337 vs 6,403 HDV/HBV_100,000_; p=0·0013). The last country to experience a significant decline in HDV incidence at a time point where the global trend was increasing is Bulgaria. Bulgaria had a significant decrease in HDV incidence from the time period of 2013-2017 to 2018-2020 (2,068 vs 718 HDV/HBV_100,000_; p=0·04) (Figure 3C).

### Identified correlations and clusters in HDV incidence in Global HDV datasets

To further characterize the international incidence of HDV over time, the 17 international HDV incidence datasets were evaluated for potential pairwise correlations (Figure 4). Hierarchal cluster analysis was performed on the pairwise correlation of the 17 datasets and identified four distinct clusters, including Cluster I (Taiwan and Macao), Cluster II (Argentina, Brazil, Germany, and Thailand), Cluster III (Bulgaria, Netherlands, New Zealand, United Kingdom, and United States), and Cluster IV (Australia, Austria, Canada, Finland, Norway, Sweden) (Figure 4A). The heatmap of the maximal HDV/HBV yearly incidence for each individual country dataset identified a temporal trend of differential timelines of peak HDV incidences across the clusters identified (Figure 4B). Additional analyses were performed based on these identified temporal clusters.

**Figure 4.**
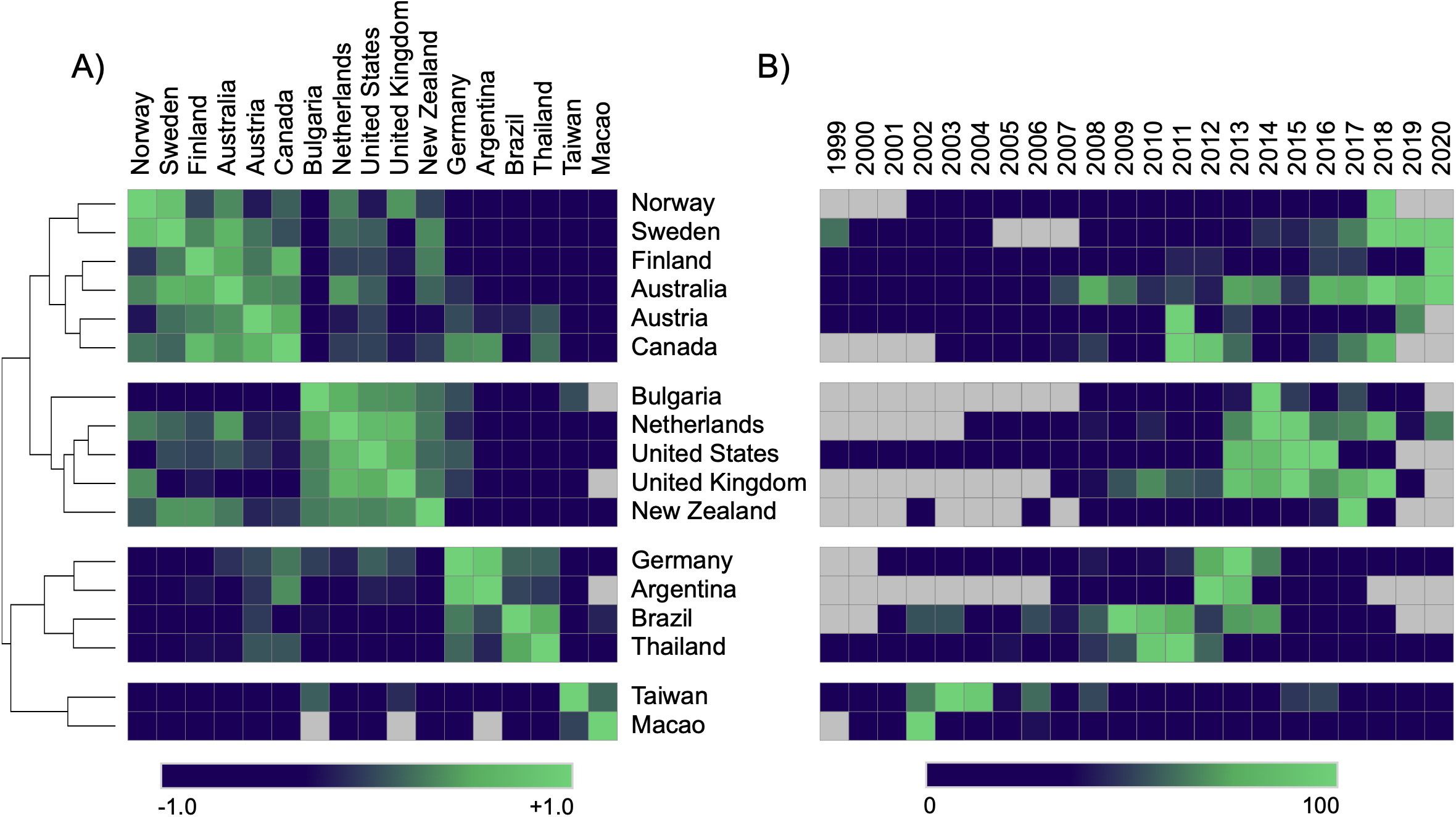
Hierarchal clustering and maximum temporal mapping among the 17 HDV/HBV yearly datasets identified distinct temporal clusters. A) Hierarchal clustering of the pairwise correlations among the 17 country datasets analyzed identified four distinct clusters: Cluster I (Macao, Taiwan), Cluster II (Argentina, Brazil, Germany, Thailand), Cluster III (Bulgaria, Netherlands, New Zealand, United Kingdom, United States) and Cluster IV (Australia, Austria, Canada, Finland, Norway, Sweden). B) Heatmap of maximal HDV incidence for each country over time series analyzed identified a temporal trend in maximal HDV/HBV incidence for each cluster.

### Significant increases in HDV incidence breakpoints for identified HDV clusters

Breakpoint analysis was performed on each temporal cluster to determine the optimal number of breaks to use for developing the best-fitting models (Supplemental Figure 4). From these best fitting models, the timelines of the structural breaks were identified (Figure 5).

**Figure 5.**
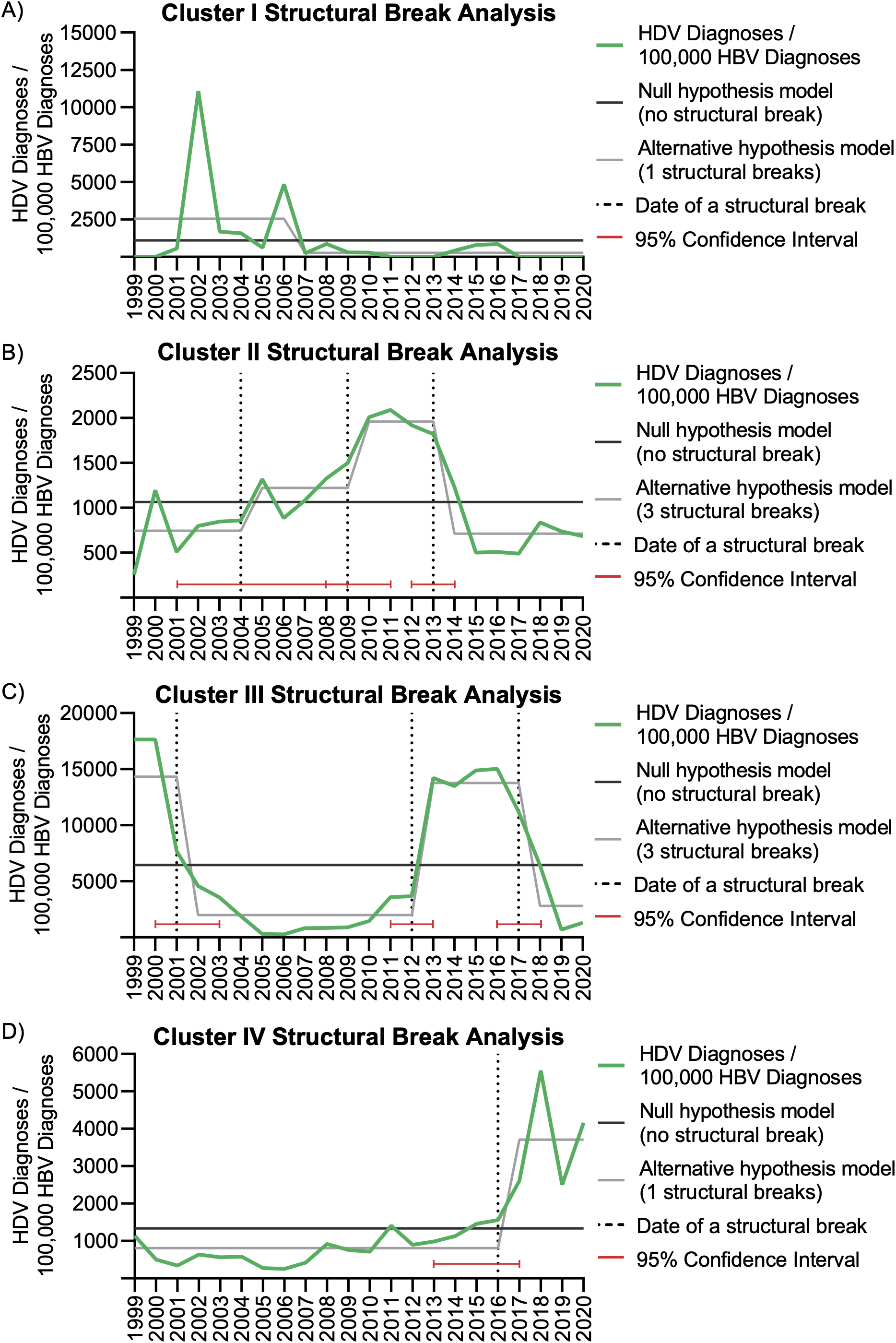
Structural Break Analysis of Four Clusters. A) No structural breaks were identified in Cluster I. B) The presence of three structural breaks in 2004, 2009, and 2013 were present in Cluster II. C) Cluster III exhibited three structural breaks in 2001, 2012 and 2017. D) A single structural break was present in Cluster IV in 2016.

In Cluster I (Macao and Taiwan), no breakpoints were identified (Figure 5A, 6A). In Cluster II (Argentina, Brazil, Germany, and Thailand), significant breakpoints were identified after 2004, 2009, and 2013 with the maximal HDV/HBV_100,000_ incidence occurring in 2010-2013 (1959, 95% CI 1775-2143 HDV/HBV_100,000_) (Figure 5B, 6B, Supplemental Table 2). In Cluster III (Bulgaria, Netherlands, New Zealand, United Kingdom, and United States), significant breakpoints were identified after 2001, 2012, and 2017, with the maximal HDV/HBV_100,000_ incidence occurring in 1999-2001 (14329, 95% CI 51-28606 HDV/HBV_100,000_), and 2013-2017 (13763, 95% CI 11825-15701 HDV/HBV_100,000_) (Figure 5C, 6C, Supplemental Table 2). In Cluster IV (Australia, Austria, Canada, Finland, Norway, and Sweden), a single significant breakpoint was identified after 2016, with the maximal HDV/HBV_100,000_ incidence occurring in 2017-2020 (1222, 95% CI 924-1520 HDV/HBV_100,000_) (Figure 5D, 6D, Supplemental Table 2).

Individual countries within the identified clusters were analyzed for significance at the determined cluster specific breakpoints (Figure 6). For Cluster II, significant increases were noted in Germany and Thailand between 2005-2009 and 2010-2013 (Germany: 1,332 vs 2,428 HDV/HBV_100,000_, FC:1·85, p=0·036; Thailand 1,630 vs 3,157 HDV/HBV_100,000_, FC:1·94, p=0·041) and significant decreases between 2010-2013 and 2014-2020 for Germany, Brazil and Thailand (Germany: 2,428 vs 1,241 HDV/HBV_100,000_, FC:-1·96, p=0·014; Brazil: 1,728 vs 615 HDV/HBV_100,000_, FC:-4·16, p=0·0007; Thailand: 3,157 vs 452 HDV/HBV_100,000_; FC:-6·98, p=0·0002) (Figure 6F). In Cluster III, significant increases were noted between 2002-2012 and 2013-2017 for Netherlands, United States, and New Zealand (Netherlands: 686 vs 1,535 HDV/HBV_100,000_, FC:2·23, p=0·0003; United States: 3,463 vs 46,901 HDV/HBV_100,000_, FC:13·54, p<0·0001; New Zealand: 1,081 vs 14,698 HDV/HBV_100,000_; FC:13·60, p=0·039). Significant decreases were present between 2013-2017 and 2018-2020 for Bulgaria (2,068 vs 718 HDV/HBV_100,000_; FC:-2·88, p=0·04) (Figure 6G). Finally, in Cluster IV, a significant decrease between 1999-2016 and 2017-2020 was present in Norway, Sweden, Finland, and Australia (Norway: 648 vs 14,629 HDV/HBV_100,000_, FC:22·58, p<0·0001; Sweden: 2,218 vs 5,896 HDV/HBV_100,000_, FC:2·66, p<0·0001; Finland: 851 vs 2,386 HDV/HBV_100,000_, FC:2·80, p=0·0068; Australia: 693 vs 1,217 HDV/HBV_100,000_, FC:1·76, p=0·0015) (Figure 6H).

**Figure 6.**
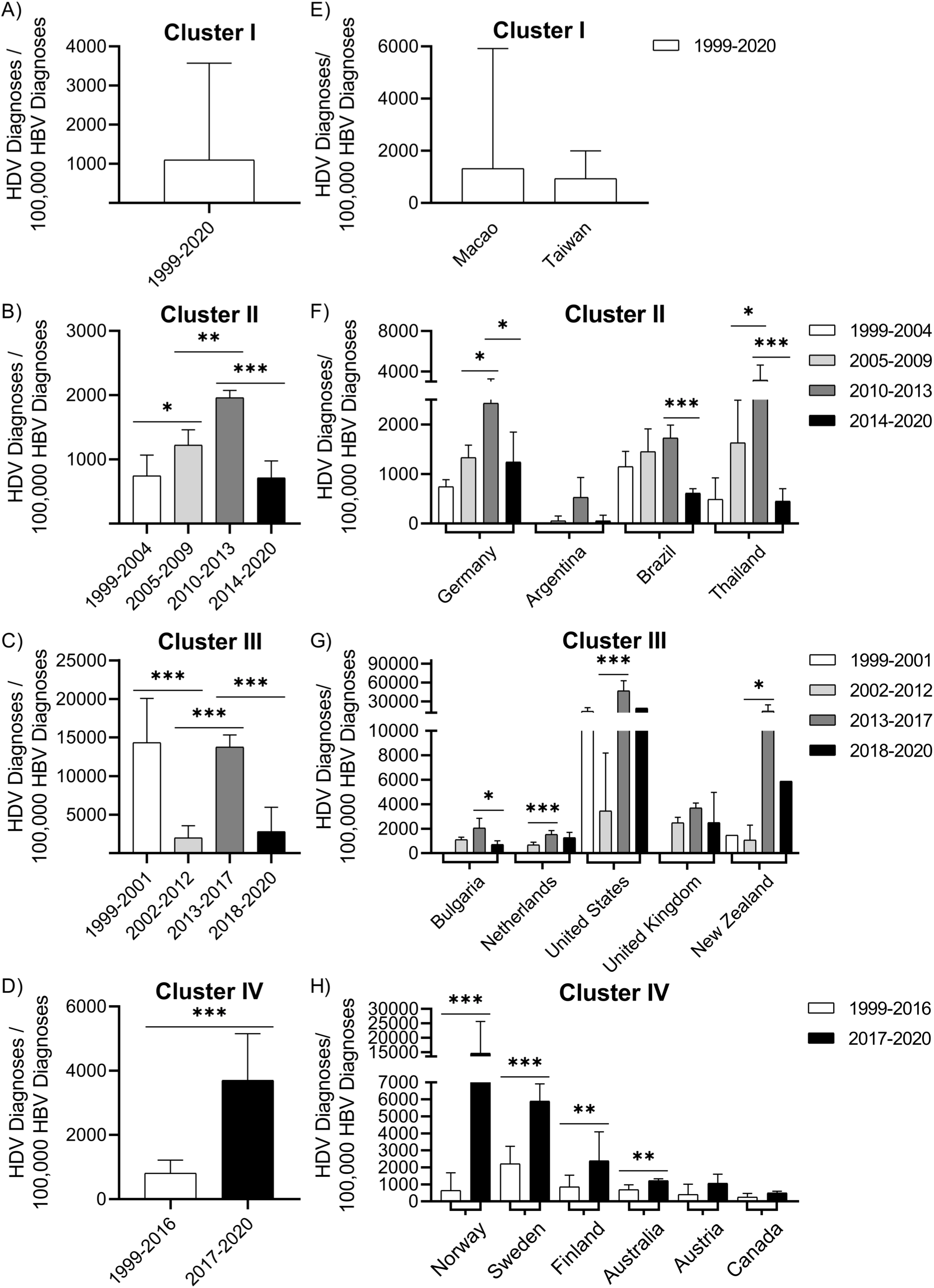
Country level HDV diagnoses per 100,000 HBV diagnoses for identified Clusters II, III and IV. A) No structural breaks and differences were identified for cluster I. B) Comparisons of years 1999-2001, 2002-2012, 2013-2017 and 2018-2020 in Cluster II. C) Comparison of 1999-2004, 2005-2009, 2010-2013 and 2014-2020 in Cluster III. D) Comparison of 1999-2016 and 2017-2020 in Cluster IV. E-H) Country-level comparisons of HDV incidence for identified clusters based on respective breakpoints. *p ≤ 0.05, **p ≤ 0.01, ***p ≤ 0.001

## DISCUSSION

Hepatitis Delta Virus (HDV) is currently defined as a rare infectious disease affecting less than 1 in 2000 people. The tracking of HDV diagnoses is attenuated by limited HDV testing of HBV-positive patient populations and the lack of active surveillance of HDV in many countries around the world. This study was developed to provide an active approach to tracking changes in HDV epidemiology as an alternative to meta-analysis of dated and restricted datasets. To date, 17 publicly accessible infectious disease datasets containing yearly incidence of HDV and HBV diagnoses have been identified and analyzed in this study. Analyses of the aggregated international HDV and HBV datasets revealed multiple recent and significant structural breaks in the timeline of HDV incidence relative to HBV diagnoses. Structural breaks were identified in the aggregated HDV time series at 2002, 2012, and 2017, with a significant increase in HDV incidence in 2013-2017 relative to all other timeframes (F=25·49; p<0·0001).

Secondary cluster analysis further identified four distinct temporal patterns of HDV/HBV diagnoses, including Cluster I (Macao and Taiwan), Cluster II (Argentina, Brazil, Germany, and Thailand), Cluster III (Bulgaria, Netherlands, New Zealand, United Kingdom, and United States), and Cluster IV (Australia, Austria, Canada, Finland, Norway, and Sweden). Further analysis of structural breaks in the time series of the four clusters identified three distinct timeframes of increased HDV incidence: 2010-2013 (Cluster II), 2013-2017 (Cluster III), and 2017-2020 (Cluster IV) (Figure 6). Classically, transmission of HDV is defined as occurring through parenteral, sexual, contaminated blood or blood products, and vertical exposure as a co-infection with HBV, or superinfection in chronic HBV carriers^1,28,29^. The observed pattern of intercontinental clusters of increased HDV incidence in the absence of a similar increase in HBV coinfections within the same population is uncharacteristic of HDV. Further analyses are required on the collective and temporal shifts in the timelines of HDV incidence to identify underlying etiology.

Our reported increase of HDV diagnoses shares a similar pattern with recent publications of HDV epidemiology. In France, a study by Servant-Delmas, et al. was conducted using viral data (1997-2011) from blood donors in the National Epidemiological Donors database. The incidence of HDV in HBV positive blood donors transitioned from 1·1% in 1997-2005, 4·2% in 2006, and 6·5% in 2010, and declined to 0·85% in 2011^30^. This transition occurred during a time period of steady decline of HBV diagnoses in blood donations. In Iran, Sayad, et al. evaluated HDV seroprevalence in HBV positive patients seen in the Liver Diseases Research Center of Kermanshah University of Medical Sciences. HDV prevalence in HBV positive patients fluctuated from 2·15% in 2004 to 1·28% in 2010, then back up to 3·47% in 2014, before declining again to 0·89% in 2016^31^. Similar patterns of HDV incidence were also reported in Cameroon by Noubissi-Jouegouo, et al. and in multiple datasets from the United States^32–34^. These reported shifts in HDV incidence mirror some of the structural breaks observed in our analyses. Together, these internationally shared temporal shifts in the incidence of HDV warrant active surveillance to monitor for emerging uncharacteristic transmission patterns of HDV.

Multiple factors may influence the perceived increase in the yearly incidence of HDV/HBV diagnoses. A majority of publicly available infectious disease datasets do not report the type of test used to detect HDV or HBV, the rate of compliance in testing HBV positive patients for HDV, or the number of tests conducted each year. Changes to the HDV testing paradigm and/or altered accessibility to the tests utilized to detect HDV antibody, antigen, and/or genomic sequence could significantly impact the incidence of reported HDV diagnoses. New methodologies for the detection of HDV have been developed and implemented in recent years that offer increased sensitivity for the detection of chronic and/or low level HDV signatures^35^. Multiple clinical studies evaluating new HDV therapies are currently underway^36–38^. This, in turn, may lead to an increase in testing and community awareness of HDV^36,37^. Together, multiple factors beyond a direct increase in HDV transmission may, in part, contribute to the detected increase in HDV diagnoses internationally.

The significant increase in HDV incidence in the CDC’s NHANES datasets may indicate widespread occurrence of HDV in asymptomatic HBV carriers. The NHANES HDV and HBV datasets are acquired biennially within the general United States population and HBV testing occurs independent of reported indicators of hepatitis. One-hundred percent of NHANES subjects that test positive for both HBV core antibodies (HBc-Ab) and surface antigens (HBsAg) are then tested for HDV antibodies. The significant increase in HDV in HBV positive subjects (2013-2016, >50% HDV+/HBV+) at 10 times the expected prevalence of HDV further suggests that the current HDV testing paradigms need to be reassessed (Figure 1). Based on the NHANES evaluation, 100% compliance in HDV testing of HBV positive populations may render an HDV incidence significantly higher than the estimated 5%. Currently, it is estimated that >200 million people worldwide are infected with HBV^39^.

Our knowledge of Hepatitis Delta Virus continues to rapidly evolve. HDV has been detected in non-hepatic tissue and associated with a disease phenotype independent of hepatitis, including the detection of HDV in salivary gland tissue of Sjogren’s syndrome patients^8^. Expression of HDV components in salivary gland tissue triggers a complete Sjogren’s syndrome phenotype, including development of autoantibodies (anti-SSA/Ro, anti-SSB/La), decreased saliva flow, and focal lymphocytic infiltration. Multiple recent publications have noted the capacity of HDV to package into non-HBV particles^40^ and at least eight new HDV-like sequences have been detected in diverse animal species, including birds, bats, deer, rodents, snakes, newts, toads, and termites^9–14^. Together, these reported deviations in basic HDV virology, associated phenotype, and the detection of new Kolmioviridae members^20^ further demonstrate the need for enhanced active surveillance of HDV to track the emergence of new epidemiological profiles, including new patterns of HDV transmission.

## Data Availability

All links to data used in study are provided in supplemental table.

## Author Contributions

All authors contributed to literature searches. BSF, EMC, MCH, JSN, and MLW collected data from publicly available datasets. BSF, EMC, SBA, and MLW performed statistical analyses. BSF, EMC, MCH, and MLW designed and drafted the figures. BSF, EMC, MLW prepared the initial manuscript draft. All authors contributed to, reviewed, and approved the final draft of the paper.

## Acknowledgements

We sincerely thank Dr. Wilson and Dr. Shankar for diligent proofreading of the manuscript. We would also like to thank the international government agencies that made their infectious disease datasets publicly accessible.

## List of Abbreviations

CDC: Center for Disease Control
NHANES: National Health and Nutritional Examination Survey
BIC: Bayesian Information Criterion

## FIGURE LEGENDS

**Supplemental Table 1. Publicly accessible datasets used in analysis of HDV and HBV diagnoses**.

**Supplemental Figure 1. Flow chart detailing testing strategy for HDV-Ab in NHANES 1999-2018 datasets**. Serum samples are tested for HBSAg, HBc-Ab and HBSAg-Ab. Only serum samples testing positive for both HBSAg and HBc-Ab are tested for HDV-Ab. Serum samples negative for HBSAg and HBc-Ab are not tested for HDV-Ab and are listed as negative in the NHANES datasets.

**Supplemental Figure 2. Results for the number of breaks from Bayesian Information Criterion (BIC) model selection analysis**. Models with increasing numbers of breakpoints are analyzed. The model with the lowest BIC value is the best fitting model. The plot indicates a model containing three structural breaks best fits the data.

**Supplemental Figure 3. Structural Break analysis of HDV/HBV (incidence) for all countries except United States (NHANES)**. A) Bayesian Information Criterion (BIC) analysis suggests a model containing a single structural break is the best fitting model. B) The presence of a single structural break was detected in 2013 in the aggregated datasets not including United States (NHANES).

**Supplemental Figure 4. Results for the number of breaks from Bayesian Information Criterion (BIC) model selection analysis for each identified cluster**. A) BIC analysis identified a model with 0 breaks best fits the countries contained within Cluster I. B) A model with 3 breaks was identified as the best model for Cluster II and C) Cluster III. D) A singular break model was identified as the best-fitting model for Cluster IV.

**Supplemental Table 2. HDV cluster analysis descriptive statistics**.

## REFERENCES

1. Wedemeyer, H. & Manns, M. P. Epidemiology, pathogenesis and management of hepatitis D: update and challenges ahead. Nat Rev Gastroenterol Hepatol 7, 31–40 (2010).

2. Miao, Z. et al. Estimating the global prevalence, disease progression and clinical outcome of hepatitis delta virus infection. J. Infect. Dis. (2019) doi:10.1093/infdis/jiz633.

3. Stockdale, A. J. et al. The global prevalence of hepatitis D virus infection: Systematic review and meta-analysis. J Hepatol 73, 523–532 (2020).

4. Chen, H.-Y. et al. Prevalence and burden of hepatitis D virus infection in the global population: a systematic review and meta-analysis. Gut 68, 512–521 (2019).

5. Gish, R. G. et al. Coinfection with hepatitis B and D: epidemiology, prevalence and disease in patients in Northern California. J. Gastroenterol. Hepatol. 28, 1521–1525 (2013).

6. Kushner, T., Serper, M. & Kaplan, D. E. Delta hepatitis within the Veterans Affairs medical system in the United States: Prevalence, risk factors, and outcomes. J. Hepatol. 63, 586–592 (2015).

7. Infectious Diseases | 2019 National Notifiable Conditions. https://www.n.cdc.gov/nndss/conditions/notifiable/2019/infectious-diseases/.

8. Weller, M. L. et al. Hepatitis Delta Virus Detected in Salivary Glands of Sjögren’s Syndrome Patients and Recapitulates a Sjögren’s Syndrome-Like Phenotype in Vivo. Pathog Immun 1, 12–40 (2016).

9. Wille, M. et al. A Divergent Hepatitis D-Like Agent in Birds. Viruses 10, (2018).

10. Chang, W.-S. et al. Novel hepatitis D-like agents in vertebrates and invertebrates. Virus Evol 5, (2019).

11. Hetzel, U. et al. Identification of a Novel Deltavirus in Boa Constrictors. MBio 10, (2019).

12. Paraskevopoulou, S. et al. Mammalian deltavirus without hepadnavirus coinfection in the neotropical rodent Proechimys semispinosus. Proceedings of the National Academy of Sciences 117, 17977–17983 (2020).

13. Iwamoto, M. et al. Identification of novel avian and mammalian deltaviruses provides new insights into deltavirus evolution. bioRxiv 2020.08.30.274571 (2021) doi:10.1101/2020.08.30.274571.

14. Bergner, L. et al. Diversification of mammalian deltaviruses by host shifting. PNAS 118, (2021).

15. Pérez-Vargas, J. et al. HDV-Like Viruses. Viruses 13, 1207 (2021).

16. Rizzetto, M. Hepatitis D: thirty years after. J. Hepatol. 50, 1043–1050 (2009).

17. Hepatitis Delta Virus. (Springer-Verlag, 2006).

18. Hughes, S. A., Wedemeyer, H. & Harrison, P. M. Hepatitis delta virus. The Lancet 378, 73–85 (2011).

19. Chang, J., Gudima, S. O., Tarn, C., Nie, X. & Taylor, J. M. Development of a Novel System To Study Hepatitis Delta Virus Genome Replication. Journal of Virology 79, 8182–8188 (2005).

20. Taxonomy History - Taxonomy - ICTV. https://talk.ictvonline.org/taxonomy/p/taxonomy-history?taxnode_id=202009293.

21. Bai, J. & Perron, P. Estimating and Testing Linear Models with Multiple Structural Changes. Econometrica 66, 47–78 (1998).

22. Bai, J. & Perron, P. Computation and Analysis of Multiple Structural Change Models. Journal of Applied Econometrics 18, 1–22 (2003).

23. Zeileis, A., Kleiber, C., Krämer, W. & Hornik, K. Testing and dating of structural changes in practice. Computational Statistics & Data Analysis 44, 109–123 (2003).

24. Ricci, A. et al. Listeria monocytogenes contamination of ready-to-eat foods and the risk for human health in the EU. EFSA Journal 16, e05134 (2018).

25. Maertens de Noordhout, C. et al. Burden of salmonellosis, campylobacteriosis and listeriosis: a time series analysis, Belgium, 2012 to 2020. Euro Surveill 22, (2017).

26. Pozza, F. et al. Impact of universal vaccination on the epidemiology of varicella in Veneto, Italy. Vaccine 29, 9480–9487 (2011).

27. Hansen, B. E. The New Econometrics of Structural Change: Dating Breaks in U.S. Labour Productivity. Journal of Economic Perspectives 15, 117–128 (2001).

28. Sagnelli, E., Sagnelli, C., Pisaturo, M., Macera, M. & Coppola, N. Epidemiology of acute and chronic hepatitis B and delta over the last 5 decades in Italy. World J Gastroenterol 20, 7635–7643 (2014).

29. Noureddin, M. & Gish, R. Hepatitis Delta: Epidemiology, Diagnosis and Management 36 Years After Discovery. Curr Gastroenterol Rep 16, (2014).

30. Servant-Delmas, A., Le Gal, F., Gallian, P., Gordien, E. & Laperche, S. Increasing prevalence of HDV/HBV infection over 15 years in France. Journal of Clinical Virology 59, 126–128 (2014).

31. Sayad, B. et al. Hepatitis D virus infection in Kermanshah, west of Iran: seroprevalence and viremic infections. Gastroenterol Hepatol Bed Bench 11, 145–152 (2018).

32. Noubissi-Jouegouo, L. et al. Evolutionary trends in the prevalence of anti-HDV antibodies among patients positive for HBsAg referred to a national laboratory in Cameroon from 2012 to 2017. BMC Research Notes 12, 1–6 (2019).

33. Martins, E. B. & Glenn, J. Prevalence of Hepatitis Delta Virus (HDV) Infection in the United States: Results from an ICD-10 Review. Gastroenterology 152, S1085 (2017).

34. Patel, E. U., Thio, C. L., Boon, D., Thomas, D. L. & Tobian, A. A. R. Prevalence of Hepatitis B and Hepatitis D Virus Infections in the United States, 2011-2016. Clin. Infect. Dis. (2019) doi:10.1093/cid/ciz001.

35. Pyne, M. T. et al. Sequencing of the Hepatitis D Virus RNA WHO International Standard. J. Clin. Virol. 90, 52–56 (2017).

36. Koh, C., Da, B. L. & Glenn, J. S. HBV/HDV Coinfection: A Challenge for Therapeutics. Clin Liver Dis 23, 557–572 (2019).

37. Deterding, K. & Wedemeyer, H. Beyond Pegylated Interferon-Alpha: New Treatments for Hepatitis Delta. AIDS Rev 21, 126–134 (2019).

38. Urban, S., Neumann-Haefelin, C. & Lampertico, P. Hepatitis D virus in 2021: virology, immunology and new treatment approaches for a difficult-to-treat disease. Gut (2021) doi:10.1136/gutjnl-2020-323888.

39. Razavi-Shearer, D. et al. Global prevalence, treatment, and prevention of hepatitis B virus infection in 2016: a modelling study. The Lancet Gastroenterology & Hepatology 3, 383–403 (2018).

40. Perez-Vargas, J. et al. Enveloped viruses distinct from HBV induce dissemination of hepatitis D virus in vivo. Nature Communications 10, 2098 (2019).

