## Supplemental Materials for "Significant Structural Breaks in the International Incidence of Hepatitis Delta Virus"

**Supplemental Table 1. Publicly accessible datasets used in analysis of HDV and HBV diagnoses.**

| Country | Website(s) |
|---|---|
| Argentina | Argentina Ministry of Health (http://www.msal.gob.ar/index.php/home/boletin-integrado-de-vigilancia) |
| | Argentina Ministry of Health (https://www.argentina.gob.ar/salud/epidemiologia/publicaciones) |
| | Argentina Ministry of Health (http://www.msal.gob.ar/images/stories/bes/graficos/0000001592cnt-2019-10_boletin-hepatitis.pdf) |
| Australia | Australian Government Department of Health (http://www9.health.gov.au/cda/source/cda-index.cfm) |
| Austria | Austrian Federal Ministry of Social Affairs, Health, Care and Consumer Protection (https://www.sozialministerium.at/Themen/Gesundheit/Uebertragbare-Krankheiten/Statistiken-und-Fallzahlen.html) |
| | Statistik Austria (https://statistik.at/web_de/nomenu/suchergebnisse/index.html?searchQuery=Jahrbuch%20der%20Gesundheitsstatistik) |
| | Statistik Austria (https://www.statistik.at/web_de/services/publikationen/4/index.html) |
| Brazil † | Brazil Health Information (TABNET) (SINANWIN) (http://tabnet.datasus.gov.br/cgi/deftohtm.exe?sinanwin/cnv/hepabr.def) |
| | Brazil Health Information (TABNET) (SINANNET) (http://tabnet.datasus.gov.br/cgi/deftohtm.exe?sinannet/cnv/hepabr.def) |
| Bulgaria | Bulgaria National Center of Infectious and Parasitic Diseases (https://ncipd.org/index.php?option=com_k2&view=item&layout=item&id=84&Itemid=1337) |
| Canada | BC Centre for Disease Control (http://www.bccdc.ca/health-professionals/data-reports/reportable-diseases-data-dashboard) |
| Finland | Finland Department of Health and Welfare (https://www.thl.fi/ttr/gen/rpt/tilastot.html) |
| Germany | Infectious Disease Epidemiology Annual Report – Robert Koch Institute (http://www.rki.de/DE/Content/Infekt/Jahrbuch/jahrbuch_node.html) |
| | SurvStat – Robert Koch Institute (https://survstat.rki.de/Content/Query/Create.aspx) |
| Macao | Macao Special Administrative Region Government Health Services (http://www.ssm.gov.mo/Portal/portal.aspx?lang=pt) |
| Netherlands | Netherlands National Institute of Public Health and Environment (https://www.rivm.nl/infectieziekten-bulletin) |
| New Zealand | New Zealand Ministry of Health – Public Health Surveillance (https://surv.esr.cri.nz/PDF_surveillance/MthSurvRpt) |
| | New Zealand Ministry of Health – Annual Surveillance Summary (https://surv.esr.cri.nz/surveillance/annual_surveillance.php) |
| Norway | Norway National Institute of Public Health (https://www.fhi.no/nettpub/smittevernveilederen/sykdommer-a-a/hepatitt-d/) |
| | Norway National Institute of Public Health – Laboratory Diagnostics (http://lab.fhi.no/Default.aspx) |
| Sweden | Swedish Public Health Agency – Hepatitis D (https://www.folkhalsomyndigheten.se/folkhalsorapportering-statistik/statistik-a-o/sjukdomsstatistik/hepatit-d) |
| | Swedish Public Health Agency – Hepatitis D (by county) (https://www.folkhalsomyndigheten.se/folkhalsorapportering-statistik/statistik-a-o/sjukdomsstatistik/hepatit-d/?t=county) |
| | Swedish Public Health Agency – Hepatitis B (https://www.folkhalsomyndigheten.se/folkhalsorapportering-statistik/statistik-a-o/sjukdomsstatistik/hepatit-b/?p=5671) |
| | Swedish Public Health Agency – Hepatitis B (by county) (https://www.folkhalsomyndigheten.se/folkhalsorapportering-statistik/statistik-a-o/sjukdomsstatistik/hepatit-b/?t=county) |
| Taiwan | Taiwan Centers for Disease Control (http://www.cdc.gov.tw/english/list.aspx?treeid=00ed75d6c887bb27&nowtreeid=c4e6b232ee7b2894) |
| | Taiwan Center for Disease Control – Hepatitis D (https://data.cdc.gov.tw/en/dataset/aagstable-acute-hepatitis-d) |
| | Taiwan Center for Disease Control – Hepatitis B (https://data.cdc.gov.tw/en/dataset/aagstable-acute-hepatitis-b) |
| | Taiwan Center for Disease Control – Disease Lookup (http://nidss.cdc.gov.tw/en/SingleDisease.aspx?dc=1&dt=3&disease=0703) |
| Thailand | Thailand Bureau of Epidemiology (http://www.boe.moph.go.th/boedb/surdata/disease.php?dcontent=old&ds=69) |
| United Kingdom | UK Health Protection Agency – Health Protection Report Archives (http://webarchive.nationalarchives.gov.uk/20140714090232/http://www.hpa.org.uk/hpr/archives/2013/news.htm) |
| | UK Government Publications (https://www.gov.uk/government/publications) |
| | UK Health Protection Report (https://www.gov.uk/government/publications/health-protection-report-volume-13-2019) |
| | UK Notification of Infectious Diseases (https://www.gov.uk/government/collections/notifications-of-infectious-diseases-noids#reports) |
| | UK Blood Born Viruses (https://www.gov.uk/government/publications/sentinel-surveillance-of-blood-borne-virus-testing-in-england-2016) |
| United States | United States Centers for Disease Control – NHANES (https://www.cdc.gov/nchs/nhanes/index.htm) |

**† For Brazil, the first link contains years 2001-2006 and the second link contains years 2007-present**

Supplemental Figure 1.

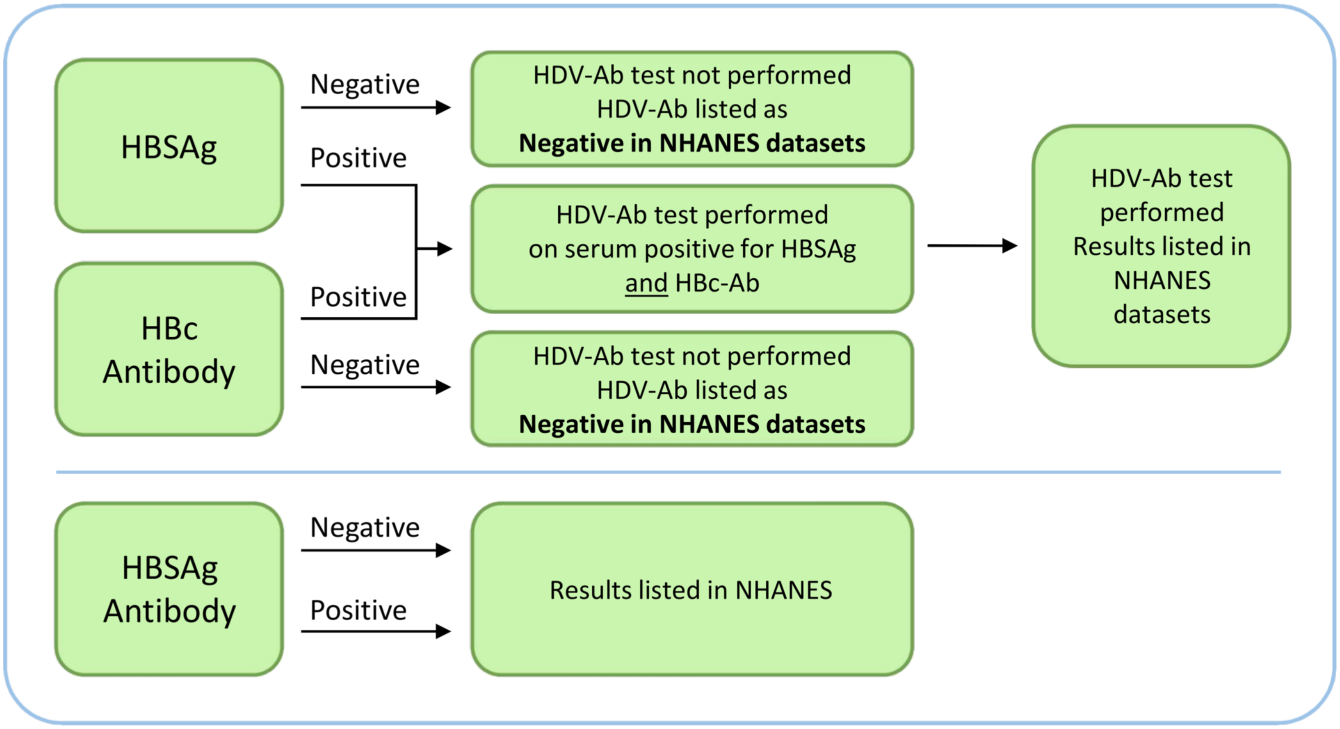

Supplemental Figure 2.

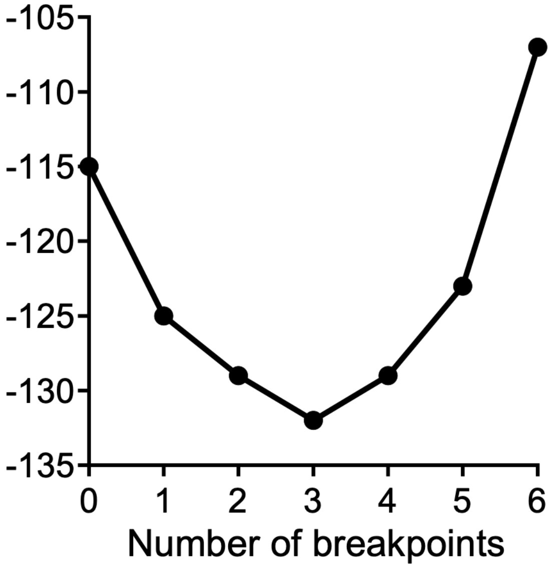

Supplemental Figure 3.

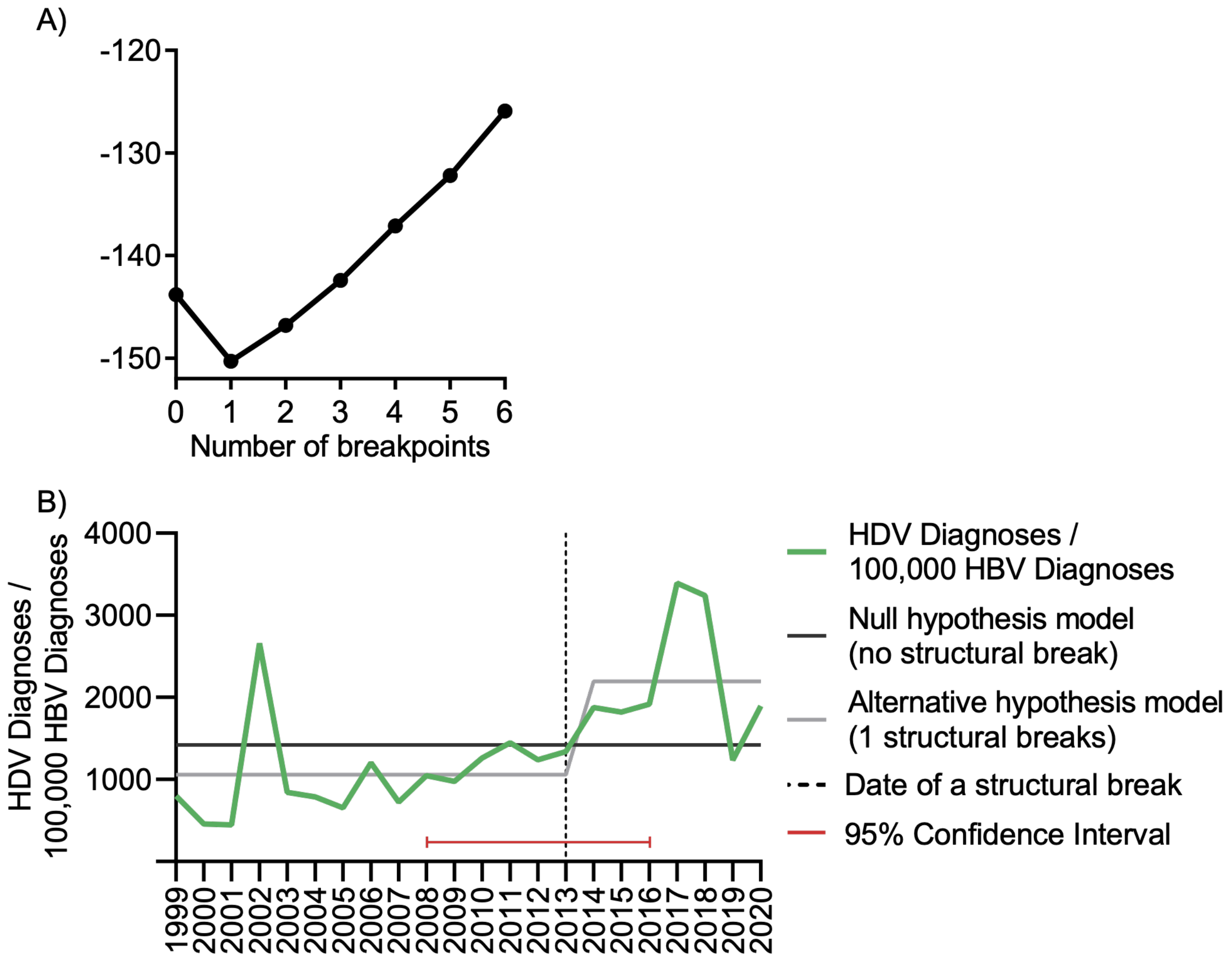

Supplemental Figure 4.

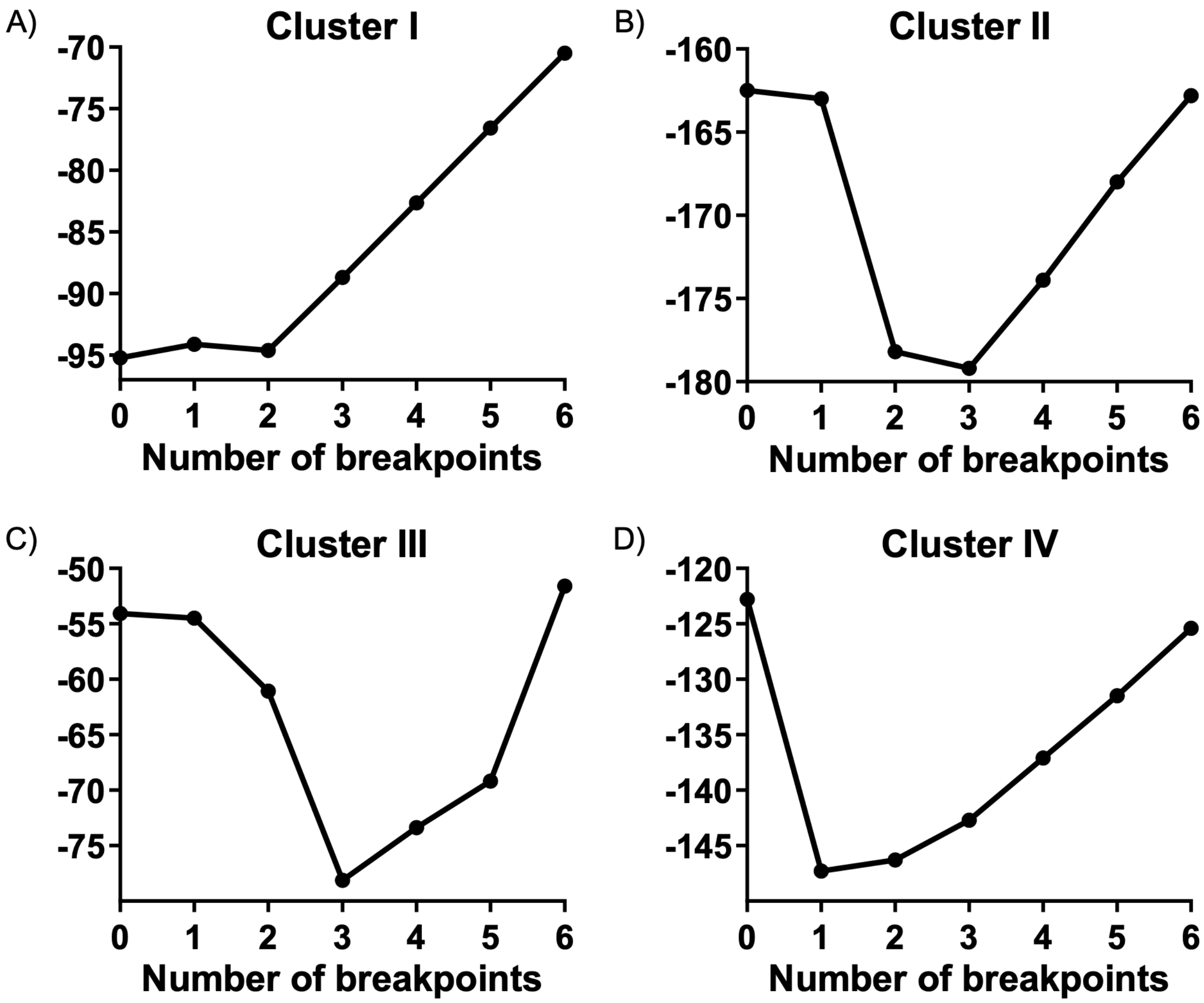

**Supplemental Table 2. Cluster HDV analysis descriptive statistics.**

| Cluster II | Mean (95% CI) HDV/$HBV_{100,000}$ | ANOVA |
|---|---|---|
| **1999-2004** | 743·9 (404·4 to 1083) | F(3,18) = 23·77 p<0·0001 |
| **2005-2009** | 1222 (923·7 to 1520) | |
| **2010-2013** | 1959 (1775 to 2143) | |
| **2014-2020** | 712·7 (468 to 957·4) | |

| Comparisons | Fold Change | P-Value |
|---|---|---|
| **1999-2004 vs. 2005-2009** | 1·64 | 0·0325 |
| **1999-2004 vs. 2010-2013** | 2·63 | <0·0001 |
| **1999-2004 vs. 2014-2020** | -1·04 | 0·9963 |
| **2005-2009 vs. 2010-2013** | 1·60 | 0·0026 |
| **2005-2009 vs. 2014-2020** | -1·71 | 0·0171 |
| **2010-2013 vs. 2014-2020** | -2·75 | <0·0001 |

| Cluster III | Mean (95% CI) HDV/$HBV_{100,000}$ | ANOVA |
|---|---|---|
| **1999-2001** | 14329 (51·48 to 28606) | F(3,18) = 35·56 p<0·0001 |
| **2002-2012** | 1992 (938 to 3045) | |
| **2013-2017** | 13763 (11825 to 15701) | |
| **2018-2020** | 2808 (-4999 to 10615) | |

| Comparisons | Fold Change | P-Value |
|---|---|---|
| **1999-2001 vs. 2002-2012** | -7·19 | <0·0001 |
| **1999-2001 vs. 2013-2017** | -1·04 | 0·9903 |
| **1999-2001 vs. 2018-2020** | -5·10 | 0·0002 |
| **2002-2012 vs. 2013-2017** | 6·91 | <0·0001 |
| **2002-2012 vs. 2018-2020** | 1·41 | 0·9614 |
| **2013-2017 vs. 2018-2020** | -4·90 | <0·0001 |

| Cluster IV | Mean (95% CI) HDV/$HBV_{100,000}$ | F-Statistic |
|---|---|---|
| **1999-2016** | 743·9 (404·4 to 1083) | F(3,17) = 12·70 p=0·0003 |
| **2017-2020** | 1222 (923·7 to 1520) | |

| Comparisons | Fold Change | P-Value |
|---|---|---|
| **1999-2016 vs. 2017-2020** | 4·59 | <0·0001 |

### Supplemental Methods

#### HDV Epidemiology in the United States (NHANES)

The National Health and Nutritional Examination Survey (NHANES) is conducted biennially by the National Centers for Health Statistics (NCHS), a part of the Centers for Disease Control and Prevention (CDC). The NHANES datasets were utilized to evaluate the passive incidence of HDV and HBV infections in the United States. The NHANES program conducts interviews and physical examinations to monitor health statistics in 5,000 people/year spanning 15 counties across the United States each year. The datasets are publicly available at https://www.cdc.gov/nchs/nhanes/. The data used in our analysis ranges from 1999/2000 to 2017/2018 and evaluates health and nutritional measurements, including the detection of antibodies to HDV (HDV-Ab), HBV surface antigens (HBSAg), HBV core antibodies (HBc-Ab), and antibodies to HBSAg (anti-HBSAg). In addition to physical examinations and clinical test results, demographic data are collected and include age at time of interview, sex, ethnicity, among other demographics. Testing for HDV and HBV in the NHANES subjects is performed in the absence of detectible hepatic symptomology that would otherwise be the primary trigger for performing tests in the healthcare setting. Detection of HDV antibodies is only performed in individuals with confirmed active HBV infection defined as having both detectible HBSAg and antibodies to HBc. Individuals that test negative for HBSAg and/or HBc antibodies are not tested for HDV antibodies and are listed as “negative” or no data is provided in the NHANES datasets.
